# Exploring the Trajectory of Catatonia in Neurodiverse and Neurotypical Pediatric Hospitalizations: A Multicenter Longitudinal Analysis

**DOI:** 10.1101/2024.06.06.24308554

**Authors:** James Luccarelli, Jacqueline A. Clauss, Tasia York, Isaac Baldwin, Simon Vandekar, Trey McGonigle, Gregory Fricchione, Catherine Fuchs, Joshua R. Smith

**Affiliations:** Harvard Medical School, Boston, MA, USA; Department of Psychiatry, Massachusetts General Hospital, Boston, MA, USA; Division of Child and Adolescent Psychiatry, Department of Psychiatry and Behavioral Sciences, Vanderbilt University Medical Center, Nashville, TN; Division of General Psychiatry, Department of Psychiatry and Behavioral Sciences; Vanderbilt University Medical Center, Nashville, TN; Department of Biostatistics, Vanderbilt University Medical Center, Nashville, TN; Vanderbilt Kennedy Center, Vanderbilt University, Nashville, TN, 37203

**Keywords:** catatonia, cohort studies, child psychiatry, neurodevelopmental disorders

## Abstract

**Objective:** Catatonia is a neuropsychiatric disorder that occurs in pediatric patients with a range of associated medical, psychiatric, and neurodevelopmental disorders (NDDs). This study describes hospital care of pediatric catatonia patients and compares treatments for neurotypical patients and those with NDDs.

**Methods:** Retrospective cohort study from 1/1/2018 to 6/1/2023 of two academic medical centers of patients aged 18 and younger with catatonia. Patients were retrospectively assessed using the clinical global impressions-improvement (CGI-I) by two independent reviewers.

**Results:** One hundred sixty-five patients were hospitalized for catatonia, of whom 50.3% had an NDD. Median age was 15. One hundred sixty-four patients were treated with a benzodiazepine, with a median maximum 24-hour dose of 6 mg lorazepam-equivalents, which did not differ for patients with and without NDDs. Electroconvulsive therapy (ECT) was utilized in 14.5% of patients. Median length of medical hospitalization was 5 days and hospitalizations were longer in neurotypical patients than in patients with NDDs. In an ordinal regression model, the probability of observing at least “much improvement” (CGI < 3) was 88.3% (95% CI: 82.4% to 92.3%), with NDD diagnosis associated with a lower odds of clinical response.

**Conclusions:** The probability of patients achieving a CGI-I score indicating at least “much improvement” was 88.3%. Administered benzodiazepine dose and ECT treatment were similar for all patients, but neurotypical patients had longer hospitalizations than those with NDDs and had a higher odds of a more favorable clinical response. Research under controlled conditions is needed to optimize and endure equitable catatonia treatment in youth.

## Introduction

Catatonia is a neuropsychiatric disorder which occurs across the lifecycle and is characterized by psychomotor disturbance, affective dysregulation, unique physical examination findings, and possible autonomic dysfunction.^1–4^ Symptoms may fluctuate rapidly during periods of illness, for instance at times presenting with muteness and stupor and at other times presenting with repetitive speech and ceaseless motion. Catatonic symptoms can be severe in nature, warranting inpatient medical care due to dehydration, poor intake of food and water, intractable aggression and hyperactivity, and autonomic instability. Children with catatonia are at a significantly elevated risk for morbidity and a 63-fold increased rate of mortality compared to same-age peers.^5^ If correctly identified, however, pediatric catatonia often responds rapidly to treatment.^6,7^ Thus, the identification and treatment of pediatric catatonia in the inpatient pediatric medical setting presents a unique opportunity for prompt and significant intervention which may drastically improve a child’s course of illness.^8–10^

Along with medically ill children, neurotypical children with psychiatric disorders and neurodiverse children are at significantly elevated risk of catatonia.^7,11–13^ For neurodiverse individuals, catatonia often presents as a deviation in baseline functioning.^14^ Common features of neurodevelopmental disorders (NDDs), including alterations of speech, eye contact, non-verbal communication, stereotypes, mannerisms, and motor function, may overlap at baseline with catatonic signs.^15,16^ In addition, the traditionally utilized Bush Francis Catatonia Rating Scale (BFCRS) is validated for neurotypical adults, and the pediatric catatonia rating scale (PCRS) is validated for neurotypical children; these scales may miss symptoms present in neurodiverse individuals with catatonia, including recurrent self-injury and loss of previously acquired skills.^17,18^ The accumulation of these challenges likely results in a delayed time to diagnosis and treatment.^19^ However, to date, we do not understand if neurodiverse children have a different longitudinal course of catatonic symptoms or if traditional first-line approaches, including benzodiazepines and electroconvulsive therapy, are equally efficacious between neurotypical and neurodiverse children.

Despite the morbidity associated with catatonic symptoms, catatonia remains understudied, especially in children. Currently, there are no comprehensive studies of the course of illness or typical treatment needed to manage catatonia in children. There also remains a significant gap in understanding how neurotypical children and children with NDD might differ in their illness course and treatment. The aim of this study is to characterize, to our knowledge, the largest sample to date of inpatient children hospitalized with pediatric catatonia. Specifically, we will examine the course, duration, and response to treatment of pediatric catatonia in the inpatient medical setting. We will then compare these outcomes between neurotypical and neurodiverse children.

## Methods

### Data Source

The clinical cohort was identified as previously described.^4^ Clinical records from two large health systems were queried for patients between 1/1/2018 and 6/1/2023 aged 18 and younger and with a discharge diagnostic code for catatonia (F06.1 or F20.2). Identified records were then manually reviewed and patients included if they had a clinical diagnosis of catatonia, as confirmed in clinical documentation and full Bush Francis Catatonia Rating Scale (BFCRS)^20^ documented at the time of initial catatonia diagnosis. This study was approved by the Institutional Review Board of each study site (Vanderbilt University IRB: 230097; Mass General Brigham IRB: 2022P000811) with a waiver of informed consent from participants.

### Data Extraction

Age, sex, race, ethnicity, and clinical diagnoses were extracted from the electronic health record. Patients were defined as having a NDD if an ICD-10-CM diagnosis under the headings F70-79 “Intellectual disabilities” or F80-89 “Pervasive and specific developmental disorders” was present.^21^ The principal reason for hospitalization was determined from the hospital discharge summary. The principal reason for hospitalization was categorized as unspecified catatonia, mood or trauma-spectrum disorders, psychotic disorders, medical diagnoses, or neurodevelopmental disorders. Length of medical hospitalization was also extracted from the discharge summary. In cases where the individual was transferred from an acute medical hospitalization to a psychiatric hospital or rehabilitation hospital and records were available for the psychiatric or rehabilitation hospitalization following transfer, the LOS for both periods were combined into an overall LOS. Initial BFCRS, discharge BFCRS (if present), maximum 24-hour benzodiazepine dosing (in milligrams of lorazepam equivalents, with 0.5 mg clonazepam, 5 mg of diazepam, and 2 mg of midazolam considered equivalent to 1 mg lorazepam), discharge benzodiazepine dosing (in milligrams of lorazepam equivalents), and the performance of electroconvulsive therapy (ECT) during hospitalization (yes/no) were extracted from the medication administration record and clinical notes.

A retrospective clinical global impressions-improvement (CGI-I)^22^ score was assigned for each patient between time of admission and time of discharge. The CGI-I score was determined independently by two raters at each study site based on review of clinical documentation of each patient, including admission records, discharge records, progress notes, and nursing notes. Each author was blinded to the results determined by their co-authors; thus two separate retrospective CGI-I scores were computed for each patient by four separate raters.

### Statistical Analysis

Demographics and diagnoses are presented using descriptive statistics, with patients with and without NDD diagnoses compared using χ^2^ and Mann–Whitney U tests. Differences in BFCRS at time of diagnosis and at discharge were compared using paired t-tests. Interrater reliability appropriate for the ordinal retrospective CGI-I scores were calculated for each site using Gwet’s AC_2._.^23^ Using CGI-I data, we fit an ordinal regression model adjusting for reviewer accounting for intra-subject correlation using robust standard errors with a working independence covariance structure.^24,25^ An additional ordinal regression model was run with age, sex, study site, index BFCRS score, and NDD diagnosis (yes/no) as independent variables. All tests were 2-sided, with a prespecified significance threshold of *p* < 0.05, without correction for multiple testing. Statistical analyses were performed using SPSS (Version 29.0. Armonk, NY: IBM Corp) and R Statistical Software (v4.2.1; R Core Team 2022). Statistical output and code for the regression analyses are included in the Supplementary Material.

## Results

In total, 165 patients met inclusion criteria, including 92 males (55.8%) and 73 females (44.2%) (Table 1). Median age was 15 years, with an interquartile range of 12 to 16. NDDs were present for 83 patients (50.3%), which included autism spectrum disorder without intellectual impairment (N=30, 36.1%), autism spectrum disorder with intellectual disability (N=28, 33.7%), intellectual disability (N=8, 9.6%), and other neurodevelopmental disorders (N=17, 20.5%).

**Table 1:** baseline demographics for pediatric patients with catatonia, overall and divided by NDD status. Listed significances compare hospitalizations for patients with NDD and those without.

|  | *Overall* |  | *NDD History* |  | *No NDD History* |  | *Significance* |
| --- | --- | --- | --- | --- | --- | --- | --- |
|  | N | % | N | % | N | % |  |
| *N* | 165 |  | 83 |  | 82 |  |  |
| *Sex* | | | | | | | $\chi^2 (1, N = 165) = 4.44, p = 0.035$ |
| *Male* | 92 | 55.8 | 53 | 63.9 | 39 | 47.6 |  |
| *Female* | 73 | 44.2 | 30 | 36.1 | 43 | 52.4 |  |
| *Age (median, IQR)* | 15 (12 to 16) | | 14 (11 to 16) | | 15 (14 to 17) | | $U = 2504, p = 0.003$ |
| < 13 | 42 | 25.5 | 28 | 33.7 | 14 | 17.1 |  |
| 13-15 | 54 | 32.7 | 26 | 31.2 | 28 | 34.1 |  |
| 16-18 | 69 | 41.8 | 29 | 34.9 | 40 | 48.8 |  |
| *Study Site* | | | | | | | $\chi^2 (1, N = 165) = 2.15, p = 0.142$ |
| *Site 1* | 136 | 82.4 | 72 | 86.7 | 64 | 78.0 |  |
| *Site 2* | 29 | 17.6 | 11 | 13.3 | 18 | 22.0 |  |
| *Race* | | | | | | | $\chi^2 (3, N = 165) = 5.91, p = 0.116$ |
| *Asian* | 6 | 3.6 | 3 | 3.6 | 3 | 3.7 |  |
| *Black* | 43 | 26.1 | 16 | 19.3 | 27 | 32.9 |  |
| *White* | 107 | 64.8 | 61 | 73.5 | 46 | 56.1 |  |
| *Other* | 9 | 5.5 | 3 | 3.6 | 6 | 7.3 |  |
| *Ethnicity* | | | | | | | $\chi^2 (1, N = 165) = 5.86, p = 0.015$ |
| *Hispanic* | 25 | 15.2 | 7 | 8.4 | 18 | 22.0 |  |
| *Not Hispanic* | 140 | 84.8 | 76 | 91.6 | 64 | 78.0 |  |
| *Primary Diagnosis* | | | | | | | $\chi^2 (4, N = 165) = 53.5, p < 0.001$ |
| *Unspecified Catatonia* | 40 | 24.2 | 26 | 31.3 | 14 | 17.1 |  |
| *Mood or Trauma Disorder* | 23 | 13.9 | 7 | 8.4 | 16 | 19.5 |  |
| *Psychotic Disorder* | 46 | 27.9 | 12 | 14.5 | 34 | 41.5 |  |
| *Medical Condition* | 25 | 15.2 | 7 | 8.4 | 18 | 22.0 |  |
| *Neurodevelopmental Disorder* | 31 | 18.8 | 31 | 37.3 | 0 | 0.0 |  |
| *Index BFCRS (median, IQR)* | 15 (11 to 20) | | 17 (11 to 21) | | 14 (11 to 18) | | $U = 2744, p = 0.031$ |

Compared to neurotypical youth with catatonia, patients with NDDs and catatonia were younger (median of 14 years vs. 15; U = 2504, *p* = 0.003), more likely to be male (63.9% vs. 47.6%; χ^2^ (1, N = 165) = 4.44, *p* = 0.035), less likely to be Hispanic (8.4% vs. 22.0%; χ^2^ (1, N = 165) = 5.86, *p* = 0.015), and had higher initial BFCRS scores (median of 17 vs. 14; U = 2744, *p* = 0.031).

One-hundred sixty-four (164, 99.4%) patients were treated with a benzodiazepine during hospitalization, with 128 (78.5%) receiving treatment with one benzodiazepine medication (lorazepam in 112, clonazepam in 14, and diazepam in 2), 30 (18.4%) receiving two different benzodiazepines during hospitalization, and 6 (3.6%) receiving three or more benzodiazepines. The number of benzodiazepines received by neurotypical children and those with NDDs was significantly different (χ^2^ (2, N = 164) = 6.57, *p* = 0.035), with 86.4% of neurotypical children treated with a single benzodiazepine compared to 69.9% of children with NDDs. The median maximum benzodiazepine dose in a 24-hour period was 6 mg of lorazepam (IQR 3 to 12), which did not differ between patients with and without NDDs (U = 3202, *p* = 0.691) (Table 2). At time of discharge 147 patients (89.1%) remained on a benzodiazepine, with a median discharge dose of 3 mg of lorazepam in a 24-hour period (IQR 1.5 to 6). The median discharge dose did not differ between patients with and without NDDs (U = 3197, *p*= 0.587). The distribution of maximum and discharge benzodiazepine doses is graphed in Figure S1.

**Table 2:**
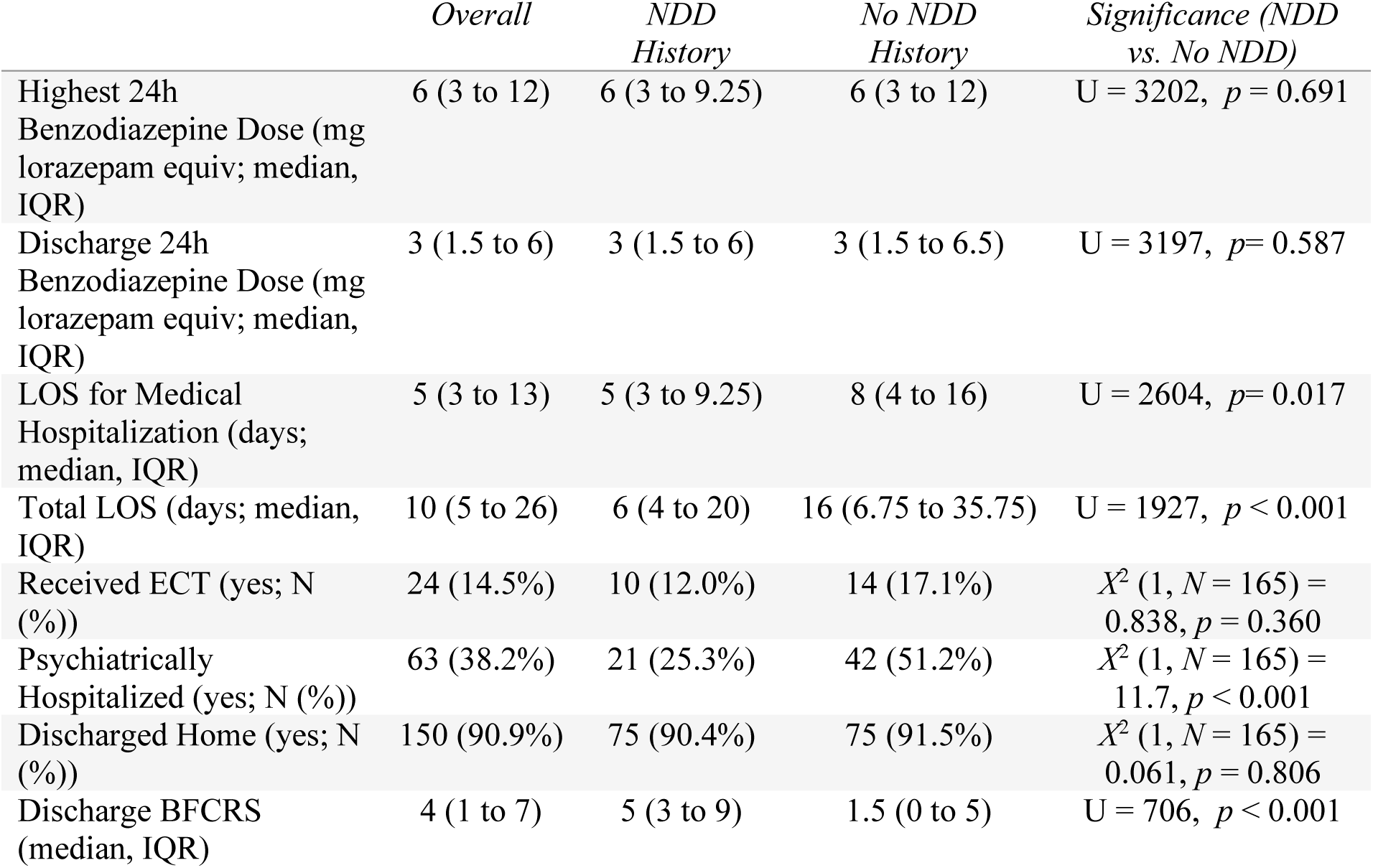
benzodiazepine dosing, hospital length of stay, ECT receipt, and discharge disease severity for pediatric catatonia hospitalizations, both overall and divided by NDD status. Listed significances compare hospitalizations for patients with NDD and those without.

Discharge BFCRS was documented for 103 patients (62.4%). Among these individuals, BFCRS decreased significantly from 16.3 ± 6.2 at baseline to 4.7 ± 4.4 at discharge (mean difference: 11.7; t(102) = 17.4; *p* < 0.001). The distribution of index and discharge BFCRS for these patients is graphed in Figure 1 and Figure S2.

**Figure 1:**
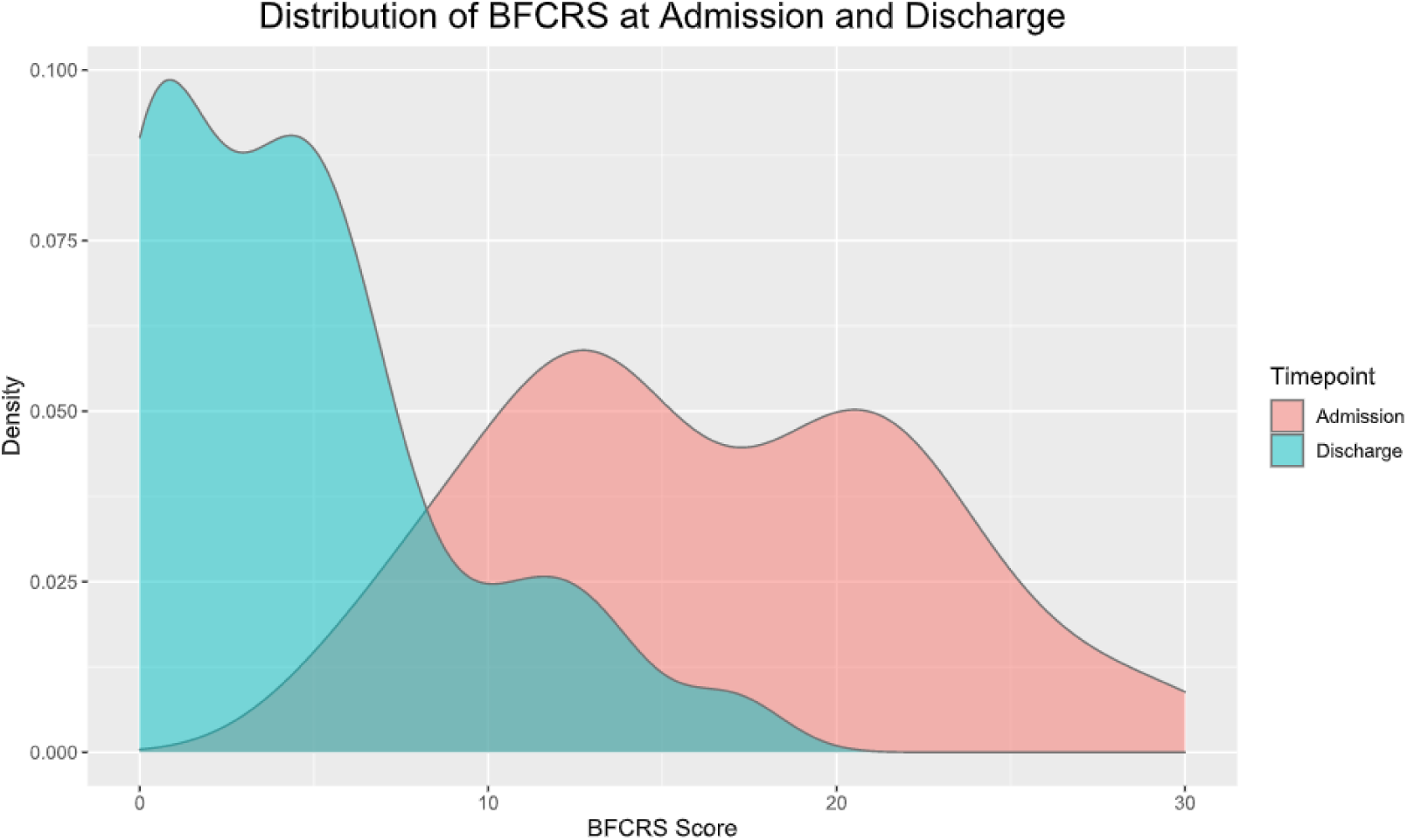
Kernel density plot of BFCRS score at admission (red) and discharge (blue) for patients with a documented discharge BFCRS (N = 103)

The median duration of medical hospitalization for all patients was 5 days, with an IQR of 3 to 13. Medical hospitalization exceeded 30 days for 9.8% of patients, with a maximum LOS of 118 days. Neurotypical children had a longer medical LOS (median of 8 days vs. 5 for those with NDD; U = 2604, *p*= 0.017). Following medical hospitalization, 63 individuals (38.2%) went on to further psychiatric hospitalization, including 51.2% of neurotypical children and 25.3% of neurodiverse children, a difference that was statistically significant (*Χ*^2^ (1, N = 165) = 11.7, p < 0.001). Length of psychiatric hospitalization was available for 55 of these patients (87.3%) and was a median of 16 days, with an IQR of 10 to 31 days. Index BFCRS was not significantly associated with length of medical hospitalization (Pearson correlation = 0.009; 95% CI: -0.145 to 0.162) or combined length of medical and psychiatric hospitalization (Pearson correlation = - 0.023; 95% CI: -0.179 to 0.135). Index BFCRS vs. medical LOS is graphed in Figure 2, and index BFCRS vs. overall LOS in Figure S3. One-hundred and fifty (90.9%) patients were discharged home after medical or psychiatric hospitalization, which was not significantly different between those with and without NDDs. There was one in-hospital death in a patient with a pediatric cancer diagnosis.

**Figure 2:**
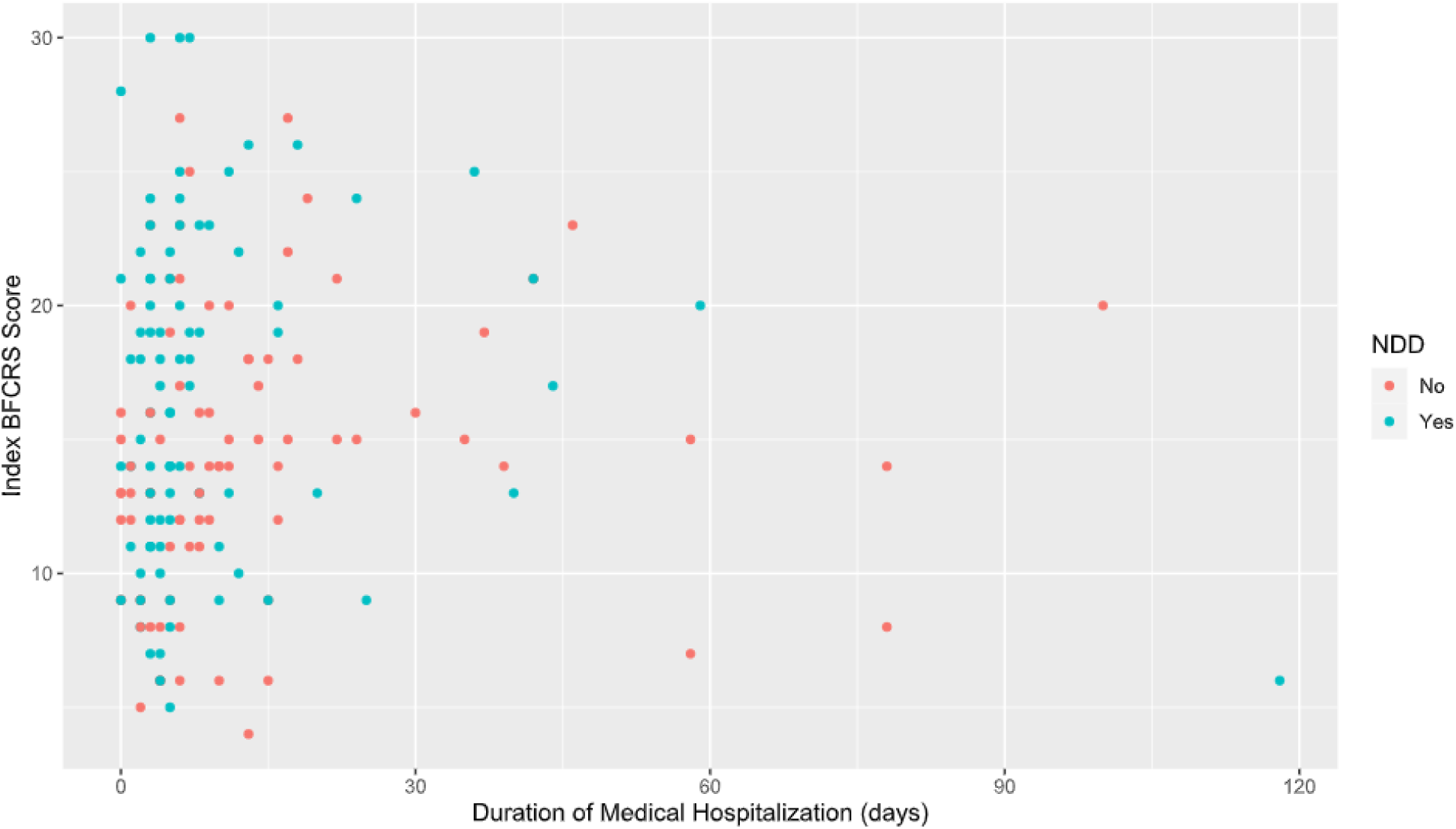
Scatterplot of initial BFCRS score vs. length of medical hospitalization in days. Patients with an NDD are colored in blue, while those without an NDD are colored red.

Overall change in illness severity between admission and discharge was quantified retrospectively for each patient using the CGI-I scale, with two reviewers independently assessing the full text of each chart. Interrater reliability for CGI-I scores was assessed using Gwet’s AC_2_ and was 0.80 (95% CI: 0.76 to 0.84) for Site 1 and 0.73 (95% CI: 0.53 to 0.94) for Site 2, indicating moderate to high correlation between reviewers. Using the CGI-I data, we fit an ordinal regression model adjusting for reviewer with robust standard errors to investigate the probability of observing improvement while accounting for intra-rater correlation. In this model, the probability of observing at least “minimal improvement” (CGI < 4) was 98.5% (95% CI: 95.4% to 99.5%), while corresponding probability of at least “much improved” (CGI < 3) was 88.3% (95% CI: 82.4% to 92.3%), and the probability of “very much improved” (CGI = 1) was 23.0% (95% CI: 17.7% to 29.2%) (Table 3). In a similar ordinal regression model with sex, study site, index BFCRS score, and NDD diagnosis (yes/no) as independent variables, the presence of a NDD diagnosis was associated with a lower odds of clinical improvement (OR 0.59; 95% CI: 0.36 to 0.95; p = 0.032), while no other variables were significantly associated with response to treatment (Table S1).

## Discussion

In this multi-site sample of 165 pediatric patients with catatonia treated within two large health systems, patients demonstrated substantial improvement in catatonia during hospitalization as measured by CGI-I scores and discharge BFCRS scores. Patients were most commonly treated with benzodiazepines, and a smaller portion were treated with ECT. Strikingly, treatment course and outcomes for neurotypical versus NDD children were different. Children with NDDs were more likely to receive more benzodiazepines and were less likely to recover fully from their illness. Additionally, children with NDDs had shorter lengths of stay, suggesting that children with NDDs may not be fully treated or may be discharged at an earlier stage in treatment.

Nearly all patients in this cohort were treated with a benzodiazepine, of which lorazepam was the most frequently utilized. The median patient had a maximum daily dosing of lorazepam of 6 mg, although there was substantial variation in this dosing. Benzodiazepines, particularly lorazepam, have demonstrated efficacy for the treatment of catatonia for more than 40 years,^26,27^ but despite this long history, there remain significant questions about optimal dosing and the overall efficacy of lorazepam for autistic patients with catatonia.^7,16,28^ A small randomized controlled crossover trial of lorazepam, dosed at 6 mg of lorazepam daily, in adult psychiatric inpatients with chronic catatonic schizophrenia failed to demonstrate a benefit from this treatment.^29^ The difference in efficacy in that trial compared to that observed here could be related to differences in weight-adjusted dosing in pediatrics, different treatment responsiveness of chronic vs. acute catatonia, diagnostic differences, or non-specific effects of general hospital treatment in patients in this sample. Systematic studies are needed to determine optimal treatment of pediatric catatonia.

Of the patients who were treated with a benzodiazepine, 21.5% of them required treatment with more than one benzodiazepine, most often longer acting agents such as clonazepam and diazepam, and of these patients, the majority of them had a diagnosis of autism spectrum disorder. Future randomized clinical trials of benzodiazepine treatment of pediatric catatonia will be required to determine optimal pharmacologic agents, dosing, and efficacy of treatment relative to placebo. By the time of discharge, most patients remained on a benzodiazepine, although at lower doses (median of 3 mg lorazepam equivalents per day); to our knowledge, there is no prospective data to support how to taper benzodiazepines in this population following discharge.

In this cohort, 14.5% of pediatric catatonia patients required treatment with ECT. This procedure has established efficacy in neurodiverse and neurotypical patients with catatonia, including those refractory to medication treatment,^6,30,31^ but remains legally restricted in many US states^32^ and with sociodemographic disparities in access.^33,34^ Barriers to access to ECT in youth have been associated with substantial harm,^35,36^ and results from this cohort point strongly to the critical need for ECT access in young patients with catatonia.

Baseline catatonia severity as measured using the BFCRS was not, however, correlated with hospital LOS nor was it associated with the odds of favorable response to treatment in an adjusted ordinal regression on CGI-I scores. The psychometric properties of the BFCRS have been explored in numerous prior studies,^37–39^ its relationship with clinical outcomes has not been investigated. These results suggest that, while the BFCRS may be an appropriate tool for identifying patients with catatonia, a higher BFCRS score may not be predictive of meaningful clinical outcomes.

Baseline rates of NDDs were high in this sample at 50.3%, and despite a higher rate of transitioning between specific benzodiazepines in the autism cohort, patients with and without NDDs did not differ substantially in benzodiazepine dosing or rate of ECT requirement. Despite these similar treatments, patients with NDDs had a lower response to treatment in a model adjusting for other baseline patient characteristics. Catatonia can be challenging to diagnose in patients with NDDs due to overlap between baseline features of such disorders and catatonic signs such as social-emotional impairment, repetitive behaviors, or impulsivity.^16,18^ Moreover, previous reports of autistic individuals with significant catatonia symptoms have demonstrated refractoriness to benzodiazepines.^7,28^ The results of our study suggest that the timely identification of catatonia in NDD patients is critical, as they may derive substantial benefit from treatment. Patients with NDDs had overall shorter length of medical hospitalization, however, and were less likely to be psychiatrically hospitalized than neurotypical youth with catatonia.

While this could be interpreted as a positive prognostic sign that individuals with NDDs require less intensive treatment, the reality is more likely that limitations in bed availability for inpatient treatment for individuals with NDD meant access was limited, and not that such patients would not benefit from additional treatment.^40^

Strengths of this study include a large sample size relative to prior publications in pediatric catatonia. Moreover, the incorporation of two geographically-distinct study sites enhances generalizability. Inclusion criteria were broad, incorporating a range of medical, psychiatric, and neurodevelopmental comorbidities. Limitations derive from the use of real-world clinical records generated as part of routine clinical care. As patients could only be included in the cohort if diagnosed with catatonia, the rate of under-diagnosis or the potential treatment responsiveness of unidentified cases of catatonia cannot be determined. Moreover, as treatments were provided as per routine care and not from a predesigned protocol, it remains unclear if patients would have benefitted from alternative treatment strategies, and so this study can only describe the treatments that were given. Additionally, the BFCRS used for catatonia assessment in this study has not been specifically validated for use in pediatric or neurodiverse patients, which should be considered when comparing data across studies. Furthermore, results from these academic health systems may not translate to other healthcare settings or to populations of different sociodemographic groups.

## Conclusion

In a multi-site retrospective cohort of 165 pediatric catatonia patients receiving inpatient treatment, there was a substantial reduction in catatonic symptom severity with treatment. Nearly all patients were treated with benzodiazepines, with ECT utilized in 14.5% of pediatric catatonia patients. Index BFCRS score did not correlate with hospital length of stay or odds of clinical improvement. Patients with NDDs had shorter hospital LOS but lower odds of clinical response in an ordinal regression model. Further research under controlled conditions is needed to optimize and ensure equitable catatonia treatment in both neurotypical and NDD youth.

## Data Availability

The data are not publicly available due to privacy restrictions from secondary use of individually identifying healthcare data; however, reasonable requests for supporting data can be directed to the corresponding author.

---

**Figure S1:**
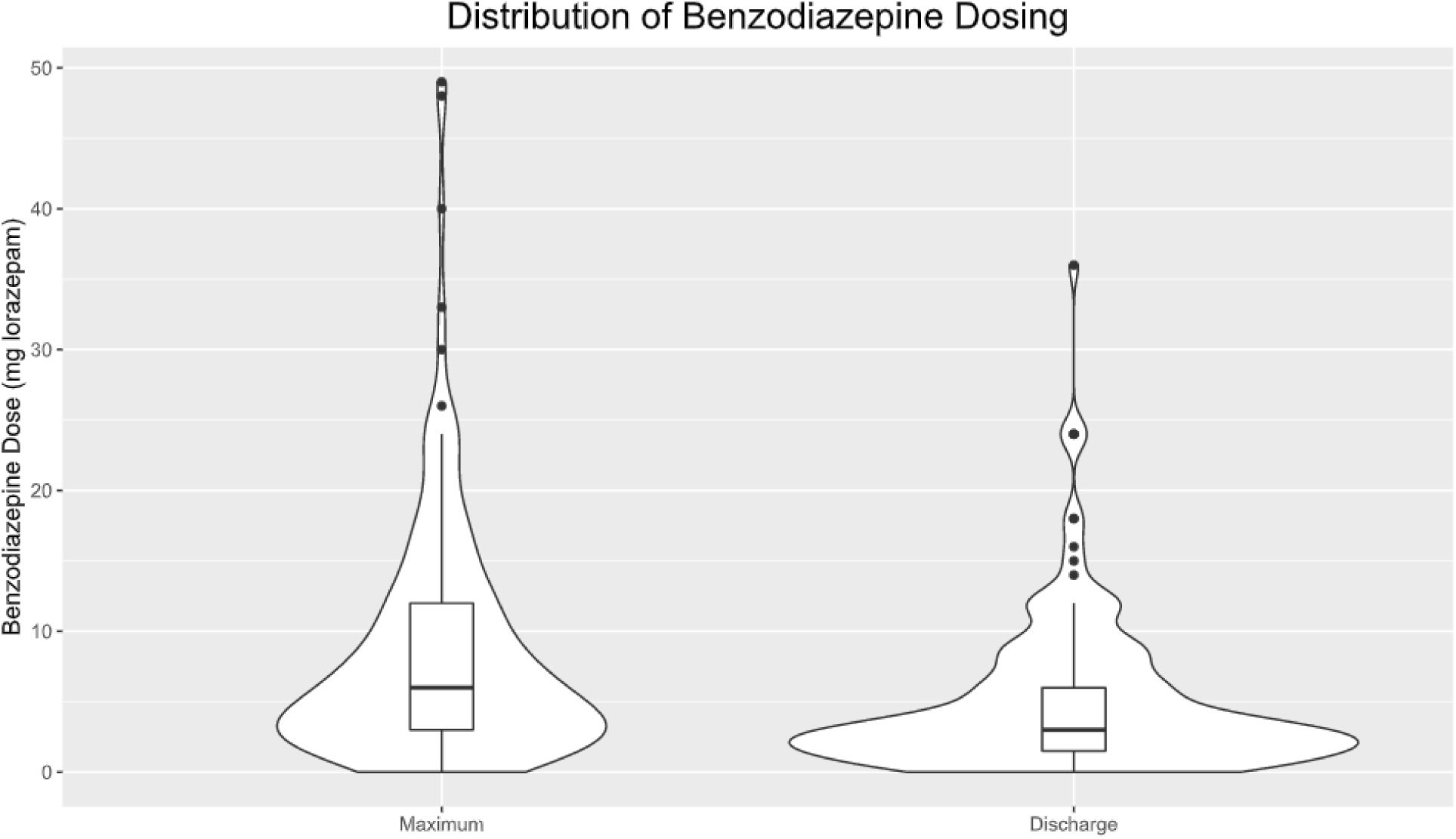
violin plots of the maximum benzodiazepine dose (left) and the discharge benzodiazepine dose (right) for pediatric patients with catatonia; the inset box in each plot displays median doses and IQR. Doses are listed in milligrams of lorazepam equivalents in a 24-hour period.

**Figure S2:**
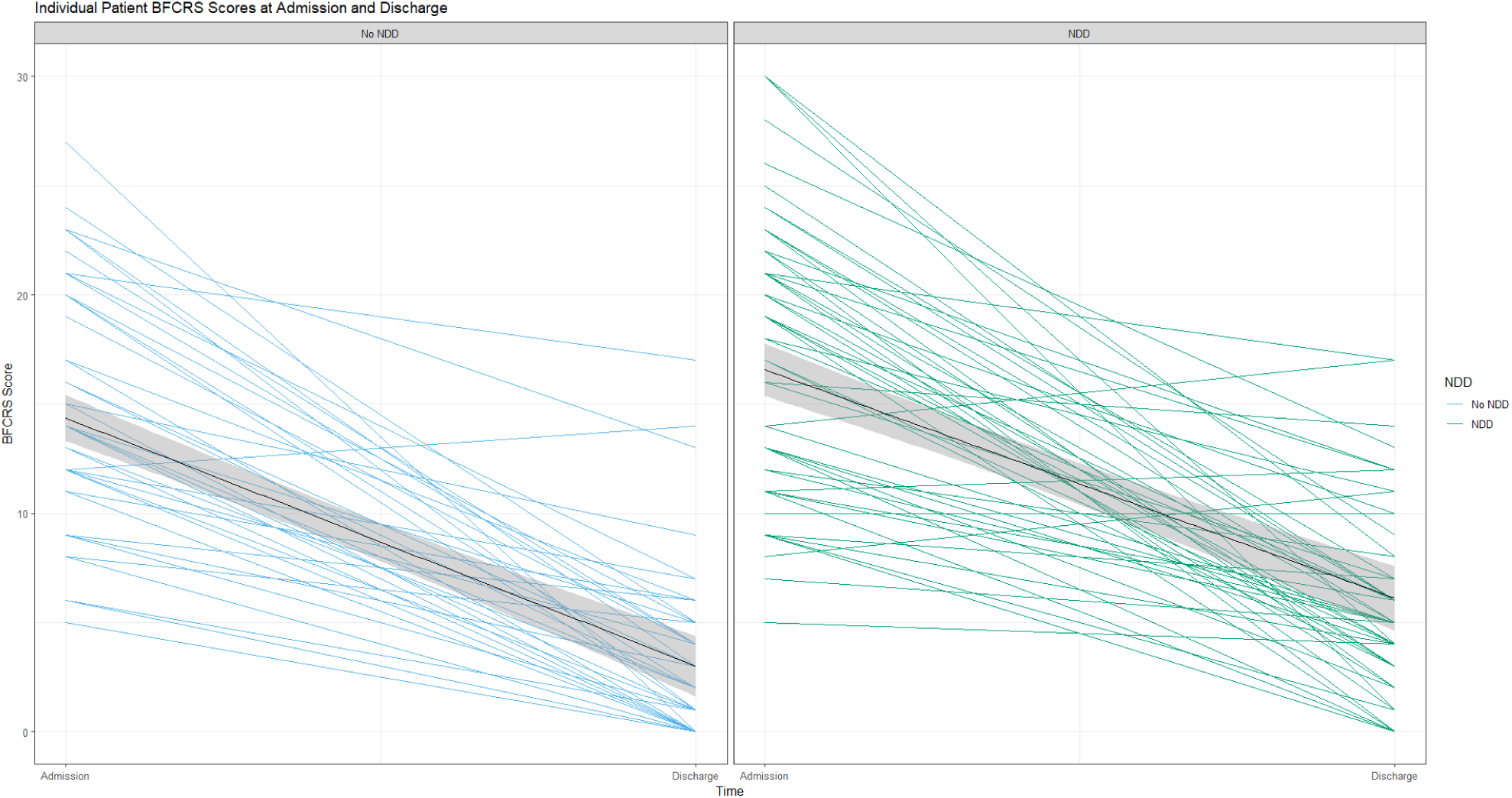
plot of admission and discharge BFCRS scores for individuals (N=103) with a documented BFCRS at time of discharge, divided by patients with NDDs (blue) and without (green). The black lines indicate the mean.

**Figure S3:**
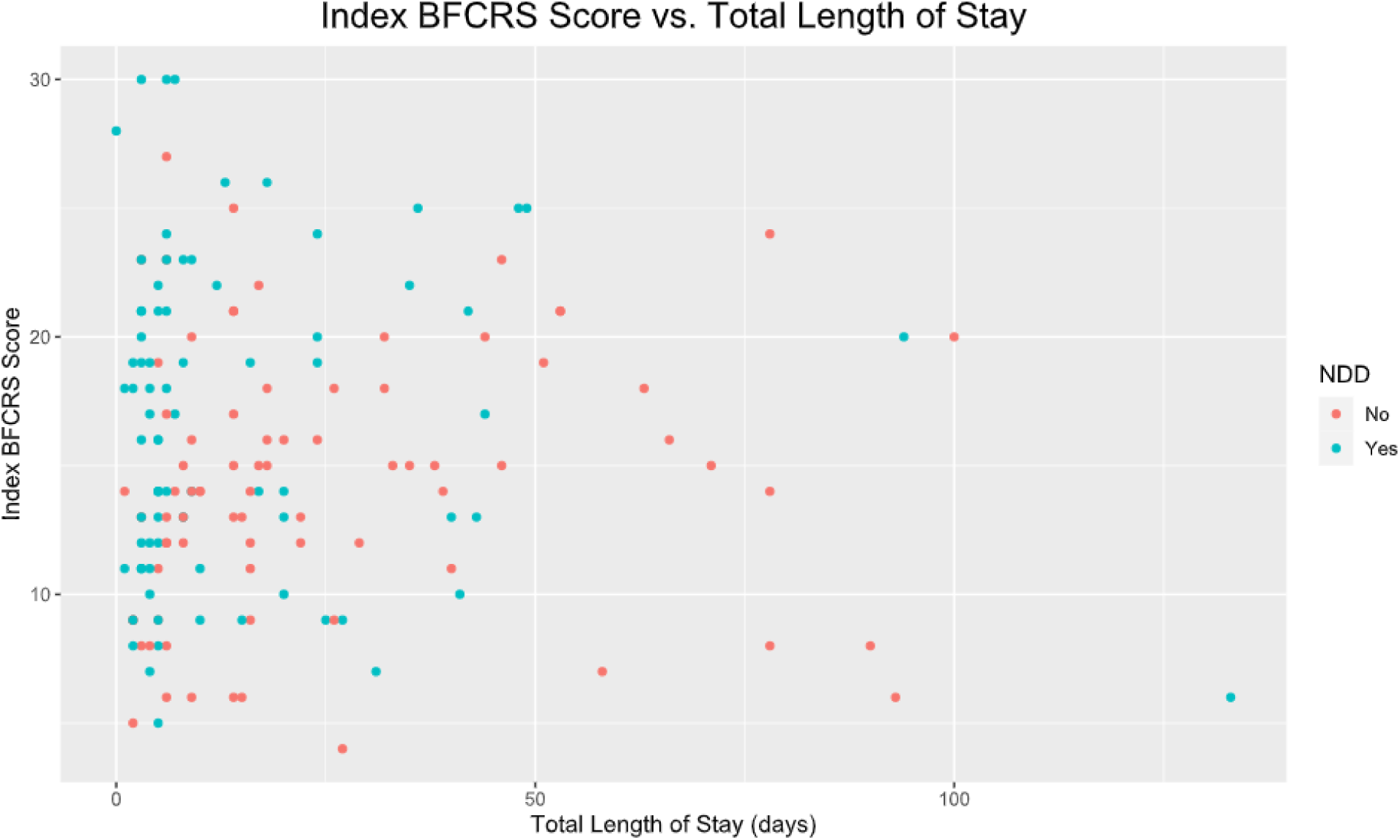
Scatterplot of initial BFCRS score vs. total length of stay (medical + psychiatric hospitalization) in days. Patients with an NDD are colored in blue, while those without an NDD are colored red.

**Table S1:**
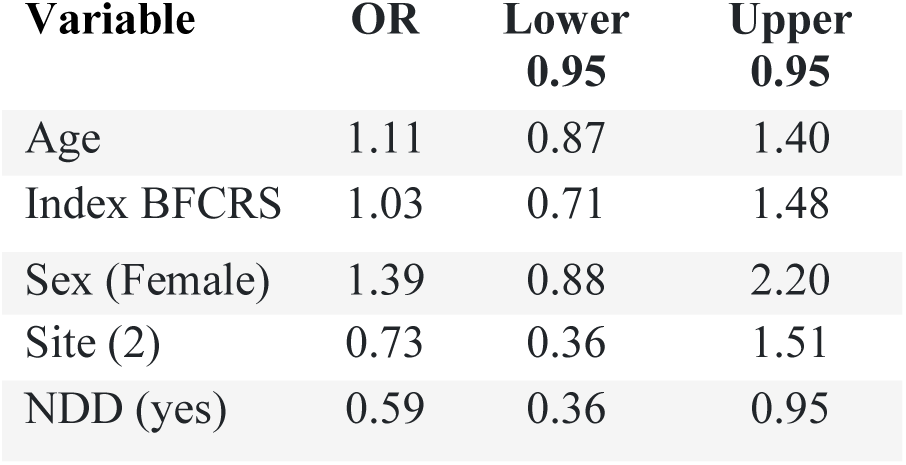
ordinal regression fit to CGI-I data adjusting for reviewer with subject-robust standard errors, with age, sex, study site, index BFCRS score, and NDD diagnosis (yes/no) as independent variables.

| **Variable** | **OR** | **Lower**<br>**0.95** | **Upper**<br>**0.95** |
| --- | --- | --- | --- |
| Age | 1.11 | 0.87 | 1.40 |
| Index BFCRS | 1.03 | 0.71 | 1.48 |
| Sex (Female) | 1.39 | 0.88 | 2.20 |
| Site (2) | 0.73 | 0.36 | 1.51 |
| NDD (yes) | 0.59 | 0.36 | 0.95 |

